# The brain mechanisms of self-identification & self-location in neurosurgical patients using virtual reality and lesion network mapping

**DOI:** 10.1101/2022.03.22.22272566

**Authors:** Sophie Betka, Julien Haemmerli, Hyeong-Dong Park, Giannina Rita Iannotti, Pavo Orepic, Eva Blondiaux, Sixto Alcoba-Banqueri, Bruno Herbelin, Christoph M. Michel, Olaf Blanke, Karl Schaller

## Abstract

**Background and Objectives:** The identification of cognitive biomarkers in preoperative counselling and their monitoring during brain surgery is of growing interest for the safe conduction of resective intracranial procedures with potential impact on the patients’ personality. The conscious experience of identifying with the body (self-identification) and of where ‘I’ am in space(self-location) are central for bodily self-consciousness (BSC). In a virtual reality (VR) paradigm using multisensory feedback, illusory self-identification and self-location over a virtual body can be induced, by manipulating the synchrony of visuo-tactile inputs. To date, no studies applied lesion network mapping (LNM) to investigate networks underlying BSC components with VR.

**Methods:** Fifteen neurosurgical patients with intra-axial and extra-axial brain lesions(8 pre-resection, 9 post-resection) performed the visuo-tactile VR paradigm. Patients subjectively rated their self-identification and self-location, after being exposed to synchronous or asynchronous visuo-tactile stimulations. We applied LNM analysis using functional data from 1015 healthy subjects and determined functional connectivity patterns related to each measure.

**Results:** In the post-surgery group, the *self-identification network* included the right inferior frontal, superior frontal and inferior temporal gyri. The *self-location network* encompassed the right parahippocampal gyrus, superior temporal gyrus, bilateral cerebellum and brainstem. No clusters survived for the pre-surgery group.

**Discussion:** Our LNM findings reveal the existence of two distinct networks for self-identification and self-location, including structures playing key roles in self-recognition or self-other distinction as well as in spatial navigation or memory processes, respectively. Such networks should be added to the portfolio of presurgical surveillance of functions related to the sense of self to improve future surgical outcomes.

**What is already known on this topic:** The development of extra- and intraoperative cognitive biomarkers is of importance for monitoring the personality of patients during cranial surgical procedures, as that may allow for individualized pre-operative counselling and intraoperative decision making. Such guidance may ultimately help to improve neuropsychological outcomes following brain surgery.

Bodily self-consciousness refers to a perceptual form of the sense of self, related to multisensory bodily inputs, which can be disrupted in neurological, psychiatric, or neurosurgical conditions.

**What this study adds:** In this study, we investigated networks associated with changes in two components bodily self-consciousness (self-identification, self-location) that we manipulated in real-time using multisensory stimulation and virtual reality in neurosurgical patients, who underwent resective surgery for brain tumors or lesions (gliomas, meningiomas, epileptic foci).

Our lesion network mapping findings reveal the existence of two distinct networks of bodily self-consciousness, including structures playing key roles in self-recognition or self-other distinction for self-identification, and in spatial navigation or memory processes for self-location.

**How this study might affect research, practice or policy:** Ideally, such networks should be identified, and their individual anatomical allocation be integrated in the surgical plan, to make them potentially amenable for functional mapping and monitoring during resective brain surgery. Ultimately, that assessment of functions related to the sense of self and personality should help.

## Introduction

In today’s intracranial resective surgery, patients’ safety relies to a large extent on the continuous analysis of electrophysiological parameters, which are obtained through intraoperative neuromonitoring (IONM)^1^. However, only selected parameters such as motor, sensory, or visual functions are routinely studied ^2^. Higher cognitive functions are not part of the standard panel of IONM, although studies have highlighted the risk of neuropsychological and personality changes after brain lesion resections^3^. The development of presurgical surveillance and of intraoperative cognitive or affective markers to drastically improve surgical outcomes has received growing interest in recent years. This also includes monitoring of the personality and sense of self of a patient during a cranial surgery^2^, highlighting the crucial need for a better understanding of the different networks underlying the sense of self.

One empirical way to test complex notions like the sense of self is to investigate bodily self-consciousness (BSC), which refers to a basic form of self-consciousness (i.e. sense of self) that is based on the simultaneous and constant integration of information coming from our body ^4^. According to classic views in consciousness science, BSC has three pillars defined as (1) self-identification which refers to the conscious experience of identifying with the body; (2) self-location which refers to where ‘I’ am in space; and (3) first-person perspective which refers to the experience of the viewpoint from where ‘I’ perceive the world ^4^. Alterations of BSC are relevant clinically and are observed in several psychiatric conditions such as depersonalization, derealization or schizophrenia as well as in neurological conditions such as epilepsy, brain tumors, or stroke ^5,6^.

After brain surgery ^2^, or in neurological conditions, such symptoms as autoscopic phenomena (i.e. altered conscious state characterized by an illusory own-body perception) characterised by the disruption of BSC and its pillars can be observed ^4,7^. For example, during heautoscopy subjects often report strong self-identification with the hallucinated duplicated own body ^8^. Moreover, heautoscopy is often associated with altered self-location and first-person perspective, and sometimes by the sensation of bi-location (i.e. the sensation of being at two places at the same time). During out-of-body experiences (OBE), patients report the feeling of being outside their physical body as well as the feeling of seeing themselves and their environment from an elevated third-person perspective ^9^, a disruption of the three BSC pillars.

However, these studies have several shortcomings. The spontaneity and paroxysmal character of such conditions as well as their rarity make the link between BSC, symptoms and potential anatomical substrates, in many cases difficult. Thus, these studies did not allow to study BSC and its alterations in real-time. Moreover, even though clinical studies have investigated the neural correlates underlying autoscopic phenomena in patients, either through case reports ^10^, intracranial brain stimulation ^9^ or quantitative lesion analyses ^7,8^, these studies did not allow investigating isolated aspects of BSC like self-identification and self-location separately.

However, research in healthy participants showed that self-identification and self-location can be modulated by experimental procedures using ambiguous or conflicting multisensory information and virtual reality (VR) ^11^. For example, in the Full-body illusion paradigm (FBI), it is possible to induce illusory self-identification and self-location over a virtual body, using multisensory feedback by manipulating the synchrony of visuo-tactile inputs, with VR^11^. Merging VR with brain imaging, brain areas such as the intraparietal sulcus, premotor cortex, temporo-parietal junction or insula have been identified as playing a key role in self-identification and self-location ^8,12–14^. However, distinct networks underlying changes in self-identification and self-location and how they are affected by damage related to primary brain lesions and neurosurgical intervention remains to be identified.

Here, we investigated networks associated with changes in self-identification and self-location during the visuo-tactile FBI paradigm in neurosurgical patients presenting intra-axial (gliomas, epileptic foci) or extra-axial tumors (meningiomas). Patients’ behavioral performances together with maps of their brain lesions were analyzed using latest lesion network mapping^15^ based on the functional connectivity of 1015 healthy subjects ^16^ to identify networks underlying changes in self-identification and self-location, during the FBI, and, thus, contributing to BSC. We hypothesized that the *self-identification network* would include structures playing a key role in self-recognition and self-other distinction such the inferior parietal lobule and lateral occipital cortex ^12–14,17^, and that the *self-location network* would be associated with regions involved in multisensory integration and spatial navigation such as temporo-parietal junction and the medial temporal structures ^17,18^.

## Materials and methods

### Patients

A total of 15 neurosurgical patients presenting intra-axial (n=1, gliomas) or extra-axial (n=4 meningiomas) tumors with a mass effect, or epileptic foci (n=1) were included from the Department of Neurosurgery, Geneva University Hospitals, Switzerland. Eight patients were tested before the resection and nine were tested after the resection of the brain lesion (pre-resection: 3 men, Age=40.5± 7.37; post-resection: 3 men, Age =53±18.87). Two patients were tested both before and after resection, with 3 months in between. Details on lesion overlap, type, lesion location, lesion size, and medication at the time of the testing for each patient are given in **Figure S3** and **Table S1**. of the supplementary section. To be included, patients needed to be over 18 and to be able to provide consent. Patients with psychiatric or neurological comorbidities or with generalized cognitive impairments were excluded. This study was approved by the ethical committee of Geneva. As part of the full protocol, the participant had to perform three tasks (their order was randomized between subjects): a full-body illusion task, a heartbeat tracking task as well as a voice recognition task. High-density EEG, cardiac activity and behavioral performances were recorded. Only the data from the full-body illusion are reported and analyzed in the present publication.

### Full-body Illusion paradigm

After taking participants’ consent to take part in the study, the instructions of the VR FBI paradigm ^11^ were explained to them. Participants were looking at an image of their own back (covered by a white towel) in a sitting position through a head-mounted display (640 × 800 resolution, 110 degrees diagonal field of view) filmed from two cameras (Logitech C510, Logitech) located two meters behind, while the experimenter applied irregular stroking on the patients’ back for 30 seconds (see **Figure 1**). During the synchronous condition, patients viewed an image of their back in real-time, whereas 0.5 s of delay was introduced during the asynchronous condition.

**Figure 1.**
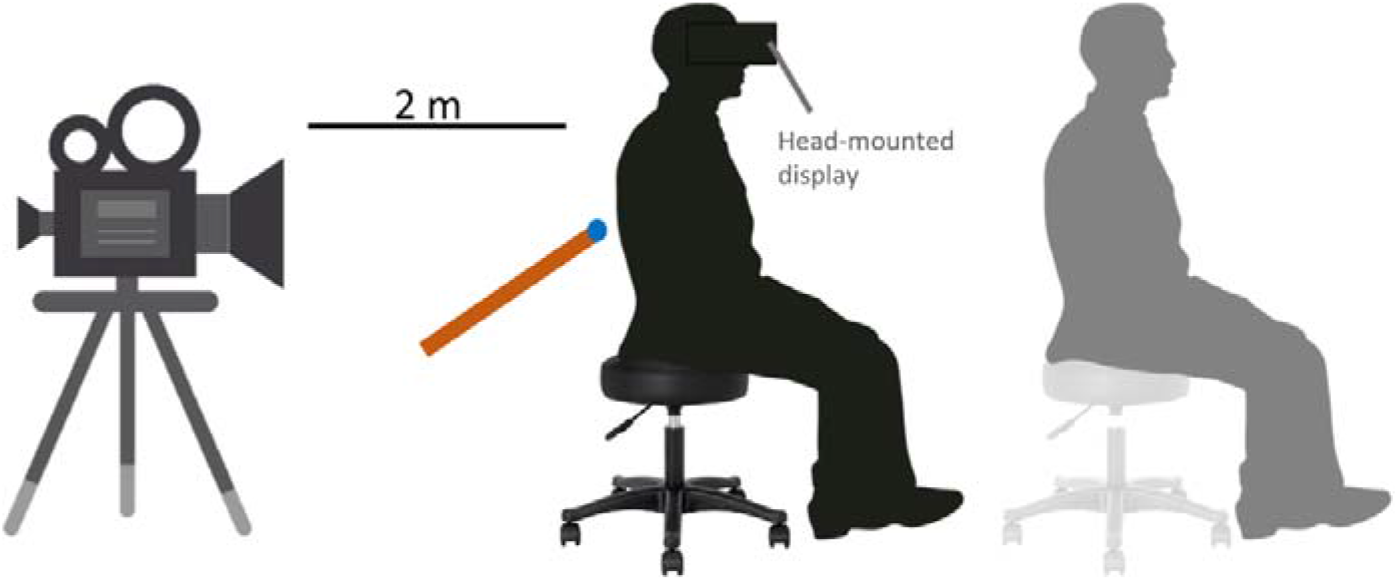
The full-body illusion (FBI) set-up. Using virtual reality, the patient (avatar in black) is wearing a VR helmet with a head-mounted display, through which a virtual avatar of that very same person can be seen from the back (avatar in light gray) and as projected at a distance of 2 m in front of the patient. Then, the experimenter applies tactile stimulations on the subject’s back using a stick and such stimulations will be displayed to the patient in the VR helmet and as seen online on the back of the avatar in VR. This is done either synchronously or asynchronously (with an additional 500-ms delay). The synchronous condition gives rise to changes in BSC, including illusory self-identification and illusory self-location over the virtual body. These alterations in BSC are evaluated using questionnaires.

The task began with a baseline and then, was composed of 4 randomized blocks (2 synchronous and 2 asynchronous; in randomized order). A mandatory break of 2 min was happening between each block.

During the baseline, participants were asked to look at a fixation cross for 60 s, then for 30s, patients viewed an image of their back being stroked (in real time or delayed based on the block’s condition) and the fixation cross appeared again for 30s. Each block was composed of five repetitions of the 30s-visuotactile stimulation period (each followed by 30s-fixation cross period). After the 5 repetitions, participants were prompted to verbally respond to the questionnaire to inquire about the state of participants’ self-identification (Q1: How strong was the feeling that the body you saw was you?), self-location (Q2: How strong was the feeling that the touch you felt was located where you saw the stroking?), and for the control purpose (Q3: I felt my body as usual, nothing changed) using a 7-point scale, ranging from 1 (bottom-extreme: Not at all) to 7 (top-extreme: Completely). We decided to use an item related to mislocation of touch as a measure of self-location^19^.

### Lesion network mapping

For each patient, we identified the lesion-derived network from each seed region of interest (ROI) following the lesion network mapping approach as described previously ^20^. The method consists of three main steps: (1) mapping each lesion into standard MNI space, (2) computing its functional connectivity at rest in a normative resting state database of healthy subjects, (3) and overlapping each of the binarized lesion-derived network together.

Given that we measured the behavioral data from each patient, we decided to modify the third step. Instead of overlapping each of the binarized lesion-derived network together, we entered the functional connectivity -not binarized-maps in a regression model, using participants scores as regressors, to highlight the networks underlying self-identification and self-location components. The analyses are explained in more detail in the following sections.

### Lesion mask creation

For each patient, depending on the nature of the lesion and the availability of the images, either one or a combination of the following sequences were used to create the lesion mask (T1-weighted, gadolinium-enhanced T1-weighted, T2-weighted, T2-weighted fluid-attenuated inversion recovery; FLAIR). Then, for each patient, images were co-registered and resliced using SPM12. The lesion was drawn on the resliced images, using the toolbox clusterize SPM toolbox and checked by qualified neurosurgeons. Finally, both lesion masks and images were normalized to the Montreal Neurological Institute space (MNI space). Lesion overlaps are presented for both groups in the supplementary section (**Figure S3**).

### MRI acquisition

For this we used the resting state and structural data from 1113 young healthy adults (age 22 to 37 years) obtained from the publicly available WU-Minn HCP 1200 Subjects Data Release from the Human connectome Project (Van Essen et al., 2012; https://db.humanconnectome.org). Scans were acquired with a using a customized 3T Connectome Scanner adapted from a Siemens Skyra (Siemens AG, Erlanger, Germany). For each participant, an anatomical image was recorded using a T1–weighted 3D MPRAGE sequence (TR = 2400 ms, TE = 2.14 ms, TI = 1000ms, Flip angle = 8°, FOV = 224×224, Voxel size = 0.7mm isotropic). For the resting sate data, a multiband gradient-echo EPI sequence was used (multiband factor = 8, 72 continuous slices, TR = 720 ms, TE = 33.1 ms, flip angle = 52°, slice thickness = 2 mm, Voxel size = 2.0 mm isotropic). Resting-state fMRI data were acquired in four runs of approximately 15 minutes each, two runs in one session and two in another session, with eyes open with relaxed fixation on a projected bright cross-hair on a dark background (and presented in a darkened room). Within each session, oblique axial acquisitions alternated between phase encoding in a right-to-left (RL) direction in one run and phase encoding in a left-to-right (LR) direction in the other run.

### Image pre-processing

We used the already pre-processed structural MRI data and pre-processed and ICA-FIX denoised resting-state fMRI data available on the HCP website. MRI preprocessing is specified in detail in ^21^. Functional data has been cleaned of structured noise through a process that pairs independent component analysis (MELODIC) with the FSL tool FIX, which automatically removes artifactual or “bad” components. For further details on this method see ^22^.

### Resting state analysis

The resting state data was analyzed using the CONN-fMRI Functional Connectivity toolbox (v.19.c, http://www.nitrc.org/projects/conn). The lesion masks were used as seed ROIs and their mean time course was extracted and correlated to all other brain voxels, limiting our analysis to voxels within the grey matter, for the 1015 subjects. In other words, a functional connectivity map was generated for each lesion, and each healthy subject, in a first-level model in CONN. Then, for each lesion, an averaged functional connectivity map was produced by averaging the 1015 functional connectivity maps, in a second-level model in CONN.

### Statistical analysis

Such averaged functional connectivity maps were entered in a second-level regression model using SPM 12. Separate models were run with each outcome as a regressor (synchronous minus asynchronous difference in self-identification & difference in self-location) and groups (pre-surgery and post-surgery), leading to a total of four models. Impact of age, gender and test-retest -e.g., if patients took part in one (pre or post) or in two sessions (pre and post)-did not significantly predict the difference (synchronous minus asynchronous) for the items (see the supplementary section, **Table S1**). Therefore, such variables were not added as covariates in the regression models. Mean and standard deviations of subjective ratings for both groups and for all conditions are also presented in the supplementary section (see **Table S3**).

For each model, an F-contrast was generated to test for brain functional activity associated with ratings. If such contrast led to significant results, we explored the direction of such association by investigating the positive (equivalent to Synchronous > Asynchronous) and negative (equivalent to Asynchronous > Synchronous) T-contrasts. Statistic images were thresholded at an initial cluster-forming threshold of p<0.001 for cluster-wise False Discovery Rate (FDR) correction for multiple comparisons at p<0.05 ^23^. Significant clusters were localized according to the Anatomy toolbox (v 2.2b ^24^).

### Data availability

All data are available from the corresponding authors upon request.

## Results

### Post-surgery group

For self-identification, functional connectivity (with the lesions as seeds) associated with ratings was examined using an F-contrast. Functional connectivity related to self-identification was observed across several brain areas such as the right inferior frontal gyrus (cluster 1; x=42 y=20 z=-14, k=168, F=69.54, p FDR-corr<0.05) and the right superior frontal gyrus (cluster 2; x= 6 y=24 z=58, k=28, F=49.10, p FDR-corr<0.05). When exploring the direction of such results using T-contrasts, we found that greater illusory self-identification (i.e. when difference between ratings for the synchronous and the asynchronous conditions are positive) was related to reduced functional connectivity between the lesions and the right inferior frontal gyrus (cluster 1; x=42 y=20 z=-14, k=297, T =8.22, p FDR-corr<0.05, **Figure *2*.A**), the right superior frontal gyrus (cluster 2; x=6 y =24 z=58, k=28, T =7.01, p FDR-corr < 0.05, **Figure *2*.B**) and the right Inferior Temporal Gyrus (cluster 3; x= 46 y =-4 z=-36, k=29, T= 6.12, p FDR-corr<0.05, **Figure *2*.C**). All activations are presented in **Table S4** of the supplementary section.

**Figure 2.**
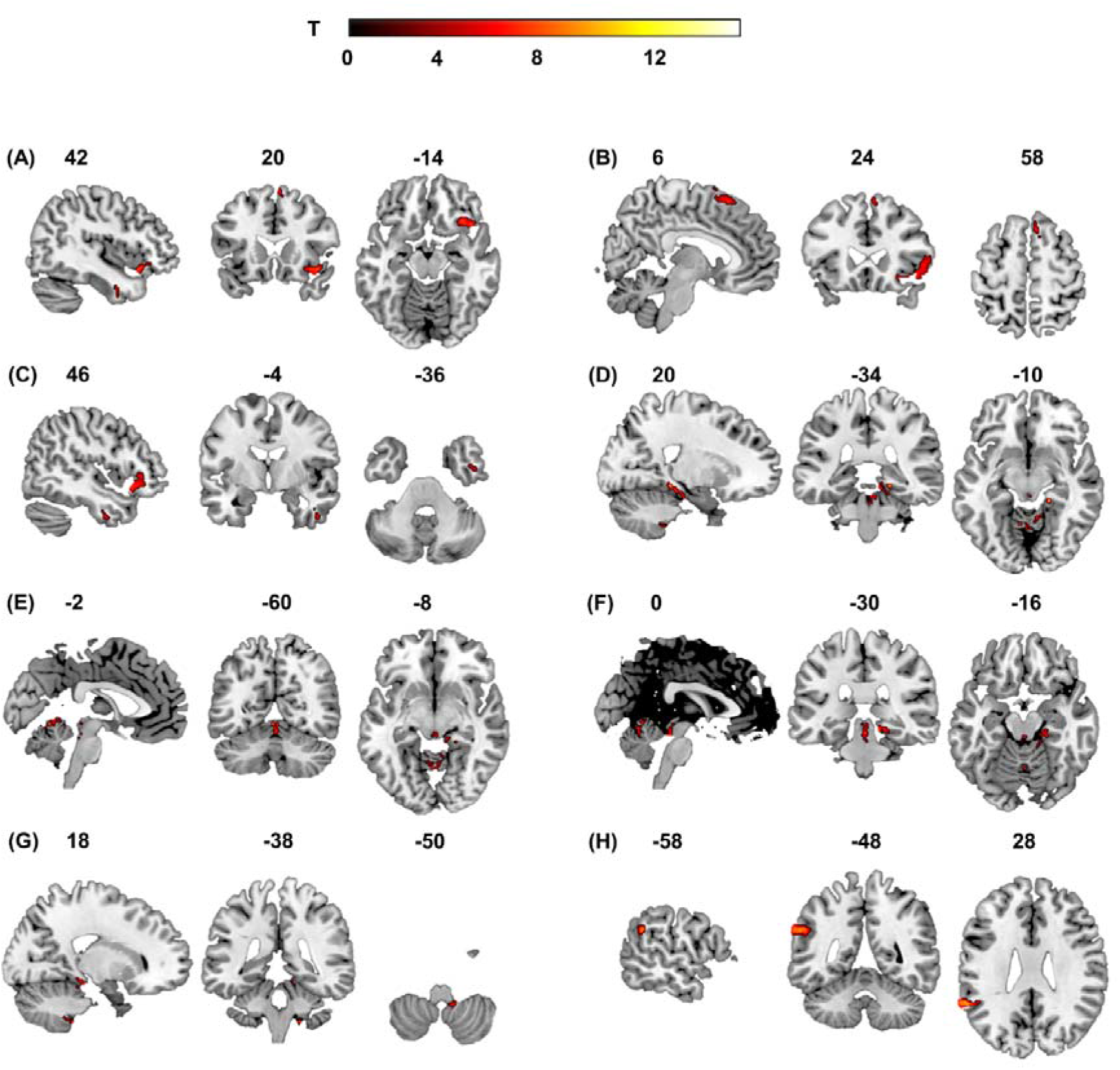
T-contrasts maps and FDR-corrected clusters for self-identification & self-location in post-surgery patients. Local maxima of significant clusters per contrast, localized according to the Anatomy toolbox (V2.2b; *Eickhoff et al*., *2005)*, in SPM12. The number above the slices indicate the x, y, z coordinates of the maximum activated voxel in standard MNI152 space. T refers to the stat at this voxel. Peaks are listed at p<0.05 FDR cluster corrected (cluster-forming threshold: p<0.001). **(A-C) Self-identification (A)** Clusters in the right temporal pole/inferior frontal gyrus when testing for the negative T-contrast testing for ratings difference equivalent to Asynch < Synch); **(B)** Clusters in the right superior frontal gyrus when testing for the negative T-contrast testing for ratings difference equivalent to Asynch < Synch); **(C)** Clusters in the right inferior temporal gyrus when testing for the negative T-contrast testing for ratings difference equivalent to Asynch < Synch); **(D-H) Self-Location (D)** Clusters in the right parahippocampal gyrus/ subiculum when testing for the positive T-contrast testing for ratings difference equivalent to Synch < Asynch); **(E)** Clusters in the bilateral cerebellum when testing for the T-contrast plus testing for ratings difference equivalent to Synch < Asynch); **(F)** Clusters in the brainstem (midbrain and pons) when testing for the positive T-plus testing for ratings difference equivalent to Synch < Asynch); **(G)** Clusters in the right cerebellum when testing for the positive T-contrast testing for ratings difference equivalent to Synch < Asynch); **(H)** Clusters in the left superior temporal gyrus when testing for the positive T-contrast testing for ratings difference equivalent to Asynch < Synch).

Applying the same approach to self-location, functional connectivity was observed across different brain areas such as the left superior temporal gyrus/temporoparietal junction (cluster 1; x=-58 y=-48 z=28, k=168, F=104.76, p FDR-corr<0.05) and the right parahippocampal gyrus/subiculum (cluster 2; x=20 y=-34 z=-10, k=44, F=161.47, p FDR-corr<0.05). When exploring the direction of such results using T-contrasts, we found that greater illusory self-location (Synch minus Asynch) was related to greater functional connectivity between the lesions, the right parahippocampal gyrus/subiculum (cluster 1; x=20 y=-34 z=-10, k=80, T= 12.71, p FDR-corr<0.05, **Figure 2.D**), the bilateral cerebellum (cluster 2; x=-2 y=-60 z=-8, k=55, T=10.62, p FDR-corr<0.05, **Figure *2*.E**), the brainstem - midbrain/pons- (cluster 3; x=0 y =-30 z=-16, k=53, T=8.08, p FDR-corr<0.05, **Figure *2*.F**) and the right cerebellum (cluster 4; x=18 y =-38 z=-50, k=48, T=9.09, p FDR-corr<0.05, **Figure *2*.G**). We also found that greater illusory self-location (Synch minus Asynch) was related to reduced functional connectivity between the lesions, the left temporal gyrus and the left supra marginal gyrus/ temporoparietal junction (cluster 1; x=-58 y =-48 z=28, k=108, T=10.24, p FDR-corr<0.05). All activations are presented in **Table S4** of the supplementary section.

Mean and standard deviations of subjective ratings for both groups and all conditions, as well as lesion overlaps, are also presented in the supplementary section (see **Table S3 & Figure S3**).

### Pre-surgery group

When testing the F-contrast, no voxel survived FDR-correction at p-value <0.05. This was observed for both self-identification and self-location.

## Discussion

The main motivation to conduct the present study was to advance understanding of the brain regions implicated in the sense of self to predict unwarranted neurosurgical side effects impacting self and personality. Here, we investigated networks associated with changes in self-identification and self-location induced experimentally by the visuo-tactile FBI paradigm in neurosurgical patients with focal cerebral lesions caused by brain lesions and epileptic foci. To do so, pre- and post-surgery patients’ behavioral performances together with maps of their brain lesions were analyzed using lesion network mapping based on the functional connectivity of 1015 healthy subjects from the Human Connectome Project. We report in the post-surgery group that the *self-identification network* includes the right inferior frontal gyrus, the right temporal pole, the right superior frontal gyrus, and the right inferior temporal gyrus. This differed for the *self-location network* which encompasses the superior temporal gyrus, the right parahippocampal gyrus, the bilateral cerebellum, the brainstem.

The involvement of the right inferior frontal gyrus in self-identification is consistent with studies showing its role in self-recognition^25^, in conflict monitoring^26^ but also in self-other distinction^27^. More recently, a lesion network analysis based on lesions from patients experiencing autoscopic phenomena showed that the right inferior frontal gyrus was part of the network underlying OBE^7^. This finding is also consistent with the involvement of the right inferior frontal gyrus in perspective taking^28^. Similarly, the right inferior temporal gyrus is known to be involved in visual self-recognition processes^29^, in scene processing^30^ and in visuo-vestibular signal integration^31^. This latter brain area has also been shown to be part of the network underlying heautoscopy^7^ and OBE-like visual illusions^32^, phenomena in which self-identification is impaired.

The analysis regarding self-location revealed the involvement of different regions, including the right parahippocampal regions. In animals, spatial representations of self and other are encoded in the hippocampus^33^. In humans, some evidence suggests the involvement the hippocampus in spatial navigation and memory but also in the construction of our sense of bodily self-location^17^, in line with recent findings showing a relationship between experimental self-location manipulations and changes in grid-cell activities in medial temporal regions^34^. Also, the manipulation of the first-person body view induces changes in episodic memory performances^35^ and also led to changes in connectivity between brain areas involved episodic memory and autonoetic consciousness such as the right hippocampus and the right parahippocampal region^36^. Moreover, the role of the parahippocampal regions has been documented in local scene processing^37^, in OBE and near-death experiences^38^. More recently, medial temporal regions, including the parahippocampal gyrus region was found to be common to networks underlying several autoscopic phenomena^7^; confirming the important implication of medial temporal structures in BSC, and in self-location processing. The network underlying self-location was also related to functional connectivity with the left superior temporal gyrus, part of the temporo-parietal junction (TPJ), compatible with the implication of the superior temporal gyrus and of the TPJ in self-location^13^, perspective taking^28^ and in heautoscopy and OBE^7,9^. Finally, clusters in the bilateral cerebellum and in the brainstem (midbrain/pons) were also found to be related to self-location processing. While the cerebellum is known to be involved in perspective taking^28^, in the establishment of maps of extrapersonal space^39^, as well as in autoscopy and OBE^7^, only little information on the contribution of the brainstem to self-location processing has been documented. This being said, the brainstem (especially midbrain and pons) contains important nuclei and relays of the vestibular systems which seem then to project to the temporoparietal areas ^40^ and therefore, may play an important role in self-location.

The current findings of the study should be considered in light of several limitations. Our pre-surgery and post-surgery groups were rather small and neurosurgical patients with focal brain functional impairment caused by brain primary lesions may have difficulties in performing complex neuropsychological tasks. Also, it is important to emphasize that our functional connectivity analysis was performed on 1015 healthy subjects, to guarantee strong results. Future work should replicate our analyses in larger clinical groups. We also note that we only observed altered lesion networks in the post-surgery group but not in the pre-surgery one; this may be related to the fact that the brain tissue is compressed by the tumor, but still functional. One could speculate that post-surgery patients are therefore a better model than pre-surgery patients for the study of self-related processes. It is also possible that pre-surgery patients may have intact areas believed to play a role in BSC and that post-surgery patients display altered components of BSC du to disconnection of important pathways by the lesion’s surgical resection^3^. Unfortunately, due to different lesion locations and sizes, pre- and post-surgery groups could not be directly compared in the present work. Further work should compare the same patients before and after the surgery to identify changes in self and/or personality by associating FBI abnormalities and lesion networks. Finally, the lesion network mapping analysis is based on inference from functional connectivity data in the healthy population. Further work should test neurosurgical patients with specific lesions in brain areas involved in our *self-identification* and *self-location* networks specifically, to confirm changes in behavior regarding each BSC component and validate our findings in a more direct manner.

To conclude, by combining the study of neurosurgical patients suffering from focal brain damage caused by tumors or epileptic foci with VR, multisensory stimulation, and the use of state-of-art functional connectivity analyses based on a large data set provided by the Human Connectome Project, we were able to highlight networks underlying two important components of BSC, that are self-identification and self-location. These findings are not only relevant for consciousness science but may inform the understanding of clinicians when confronted with patients with complex symptoms including changes in self and personality, as often observed after brain surgeries^2^. Also, it is often difficult to disentangle psychological responses of patients due to a brain primary lesion or brain surgery *per se* from neurological effects due to interference with specific neural networks involved in self and personality. The highlighted networks should be added to the portfolio of pre-surgical evaluations and intra-operative surveillance of functions to improve future surgical outcomes.

## Data Availability

All data produced in the present study are available upon reasonable request to the authors

## Acknowledgements

Data were provided [in part] by the Human Connectome Project, WU-Minn Consortium (Principal Investigators: David Van Essen and Kamil Ugurbil; 1U54MH091657) funded by the 16 NIH Institutes and Centers that support the NIH Blueprint for Neuroscience Research; and by the McDonnell Center for Systems Neuroscience at Washington University. We would like to thank the staff from SCITAS who was very helpful regarding the handling the calculations on the cluster.

