## Supplementary material for "The brain mechanisms of self-identification & self-location in neurosurgical patients using virtual reality and lesion network mapping"

**Co-last authors

Author affiliations:

^1^Laboratory of Neurocognitive Science, Center for Neuroprosthetics and Brain Mind Institute, Ecole Polytechnique Fédérale de Lausanne (EPFL), Geneva, Switzerland;

^2^Department of Neurosurgery, Geneva University Hospitals & Faculty of Medicine, University of Geneva, Switzerland;

^3^Functional Brain Mapping Laboratory, Department of Fundamental Neurosciences, University Geneva, Switzerland;

^4^Department of Clinical Neurosciences, Geneva University Medical Center & Faculty of Medicine, University of Geneva, Switzerland.

Correspondence to: Dr Betka Sophie

Full address Campus Biotech H4 Chemin des Mines 9 CH-1202 Genève, Switzerland

**Running title**: Neural correlates of Bodily self-consciousness

### Supplementary material

|  | | | **Pre-surgery Group** | | |
| --- | --- | --- | --- | --- | --- |
| **Patients** | **Age Range*** | **Gender** | **Lesion (type, stade, localisation, side)** | **Size**** | **Medication** |
| Patient01 | 20 | M | Oligodendroglioma WHO Grad II, right fronto-opercular and insular location | 11583 | Leveiracépam 750x2, lacosamide 500 2x,  valproate x3 |
| Patient02 | 40 | F | Low grad glioma left frontal at the level of the posterior F1 | 890 | No Antiepileptic |
| Patient03 | 30 | M | Astrocytoma WHO Grad II right frontal, F2 | 489 | No antiepileptic |
| Patient04 | 60 | F | Astrocytoma WHO Grad II, left frontal at the level of the medial frontal gyrus | 1180 | No Antiepileptic |
| Patient05 | 50 | F | Transitional Meningioma WHO Grad I, right frontal parasagittal located | 5278 | No Antiepileptic |
| Patient06 | 50 | F | Atypical Meningioma WHO Grad II, left precentral parasagittal located | 2884 | Dexamethasone |
| Patient07 | 20 | M | Ganglioglioma WHO Grad I, right temporal at the level of T1 | 76 | Vimpat 200 mg 2x, |
| Patient08 | 10 | F | Low grad glioma involving the right hippocampus | 647 | No Antiepileptic |
|  | | | **Post-surgery Group** | | |
| **Patient** | **Age Range*** | **Gender** | **Lesion (type, stade, localisation, side)** | **Size**** | **Medication** |
| Patient09 | 50 | F | Oligoastrocytoma WHO Grad II, frontal posterior located | 24853 | Lamotrigin 200 x3, Vimpat 100 1x |
| Patient10 | 80 | F | meningioma WHO Grad I left frontal | 543 | No antiepileptic |
| Patient11 | 30 | M | Pharmaco-resistant epilepsy right parietal at the level of the precuneus | 776 | Prebabalin 150 1x, vimpat 100 2x, Vimpat 200 1x, Clonazepam if needed |
| Patient12 | 40 | F | Oligoastrocytoma WHO Grad II, right fronto-temporo-insular | 11316 | No Antiepileptic |
| Patient04 | 60 | F | Astrocytoma WHO Grad II, left frontal at the level of the medial frontal gyrus | 2554 | No Antiepileptic |
| Patient07 | 20 | M | Ganglioglioma WHO Grad I, right temporal at the level of T1 | 2529 | Vimpat 200 mg 2x, |
| Patient13 | 60 | M | Astrocytoma WHO Grad II, right temporal, recurrence post surgery | 7288 | No Antiepileptic |
| Patient14 | 50 | F | Meningioma WHO Grad I left frontal with extension to the right side through the superior sinus | 132 | No Antiepileptic |
| Patient15 | 40 | F | Anaplasic astrocytoma WHO Grad III, left temporo-posterior with extension to the left hippocampus | 12734 | Levetiracepam 250mg 2x, dropéridol 0.5, Dexamethasone |

**Table S1: Details regarding the lesion and the medication of all patients. * 10-year age range; ** in voxels**

| **Self-Identification**  **Difference (Synch - Asynch)** | **Estimate** | **Std error** | **t-value** | **p-value** |
| --- | --- | --- | --- | --- |
| Intercept | 0.708 | 1.233 | 0.574 | 0.575 |
| Gender | 0.432 | 1.046 | 0.414 | 0.686 |
| Age | -0.003 | 0.027 | -0.126 | 0.902 |
| Test-retest | -0.523 | 1.36 | -0.385 | 0.707 |
| **Self-Location**  **Difference (Synch - Asynch)** | **Estimate** | **Std error** | **t-value** | **p-value** |
| Intercept | 3.044 | 1.341 | 2.271 | 0.041 |
| Gender | 1.159 | 1.138 | 1.019 | 0.327 |
| Age | -0.048 | 0.03 | -1.637 | 0.126 |
| Test-retest | -0.979 | 1.479 | -0.662 | 0.52 |

**Table S2: Impact of age, gender and test-retest -e.g., if patients took part in one (pre or post) or in two sessions (pre and post)- on the difference in self-identification and self-location.** As age, gender and test-rested did not have a significant impact on the difference (synchronous – asynchronous) for the items, we did not include such variables in our statistical regression models.

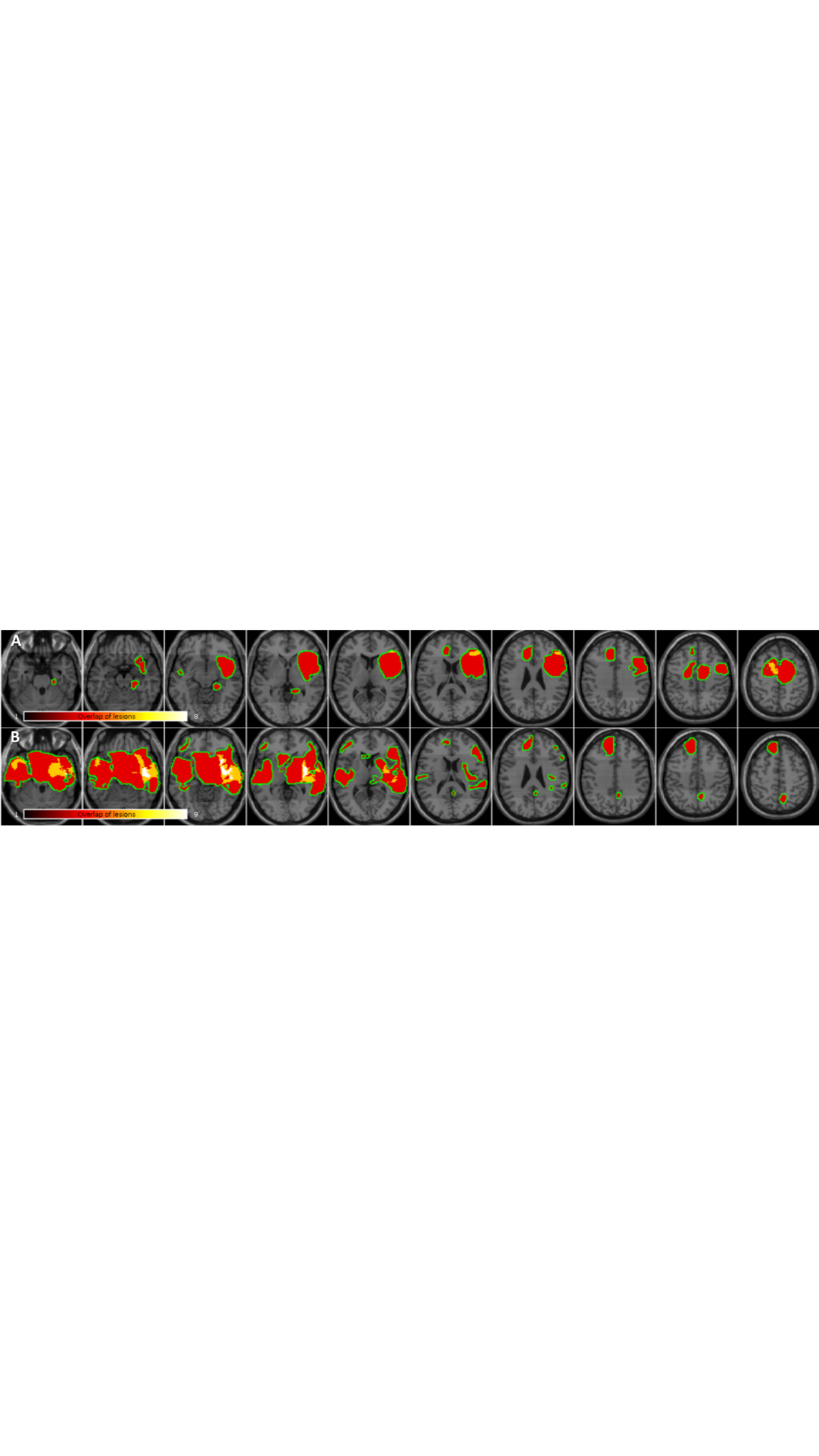

**Figure S3: Overlap of patients’ lesions for pre-surgery (n=8) (A) and post-surgery (N=9) (B) groups.**

|  | **Pre-Surgery (n=8)** | | **Post-Surgery (n=9)** | |
| --- | --- | --- | --- | --- |
|  | **Asynchronous** | **Synchronous** | **Asynchronous** | **Synchronous** |
| **Self-identification** | 5.063 (± 1.266) | 6 (±1.254) | 5.056 (± 2.171) | 5.667 (±2.062) |
| **Self-location** | 4.188 (± 1.792) | 6 (± 0.926) | 5.278 (± 1.660) | 6.278 (±0.939) |
| **Control** | 6.313(± 1.033) | 6.313(± 1.193) | 6.889 (± 0.334) | 7 (±0) |

**Table S3 Mean and standards deviations of subjective ratings for both groups and for all conditions.**

| **Self-Identification (F test)** | | | | | |
| --- | --- | --- | --- | --- | --- |
| **Cluster 1 (168 vox): F ( F> 29.25)** | **F/t** | **x** | **y** | **z** | **Area** |
| Maximum 01 | F = 69.54 | 42 | 20 | -14 | R Temporal Pole |
| Maximum 02 | F = 67.63 | 34 | 24 | -12 | R IFG (p. Orbitalis) |
| Maximum 03 | F = 67.34 | 44 | 24 | -10 | R IFG (p. Orbitalis) |
| Maximum 04 | F = 44.45 | 46 | 30 | 2 | R IFG (p. Orbitalis) |
| Maximum 05 | F = 43.23 | 50 | 28 | -6 | R IFG (p. Orbitalis) |
| Maximum 06 | F = 43.09 | 50 | 26 | -2 | R IFG (p. Orbitalis) |
| Maximum 07 | F = 42.59 | 52 | 24 | 0 | R IFG (p. Orbitalis) |
| Maximum 08 | F = 37.20 | 42 | 32 | -8 | R IFG (p. Orbitalis) |
| Maximum 09 | F = 36.02 | 54 | 26 | 6 | R IFG (p. Triangularis) |
| **Cluster 2 (28 vox): F ( F> 29.25)** |  | | | | |
| Maximum 01 | F = 49.10 | 6 | 24 | 58 | R Superior Frontal Gyrus |
| Maximum 02 | F = 40.46 | 4 | 16 | 64 | R Posterior-Medial Frontal |
| **Self-Identification (negative T test: Asynch < Synch)** | | | | | |
| **Cluster 1 (297 vox): T- ( T> 4.79)** | **F/t** | **x** | **y** | **z** | **Area** |
| Maximum 01 | T = 8.34 | 42 | 20 | -14 | R Temporal Pole |
| Maximum 02 | T = 8.22 | 34 | 24 | -12 | R IFG (p. Orbitalis) |
| Maximum 03 | T = 8.21 | 44 | 24 | -10 | R IFG (p. Orbitalis) |
| Maximum 04 | T = 6.67 | 46 | 30 | 2 | R IFG (p. Orbitalis) |
| Maximum 05 | T = 6.57 | 50 | 28 | -6 | R IFG (p. Orbitalis) |
| Maximum 06 | T = 6.56 | 50 | 26 | -2 | R IFG (p. Orbitalis) |
| Maximum 07 | T = 6.53 | 52 | 24 | 0 | R IFG (p. Orbitalis) |
| Maximum 08 | T = 6.31 | 54 | 22 | -8 | R IFG (p. Orbitalis) |
| Maximum 09 | T = 6.10 | 42 | 32 | -8 | R IFG (p. Orbitalis) |
| Maximum 10 | T = 6.00 | 54 | 26 | 6 | R IFG (p. Triangularis) |
| **Cluster 2 (62 vox): T- ( T> 4.79)** |  | | | | |
| Maximum 01 | T = 7.01 | 6 | 24 | 58 | R Superior Frontal Gyrus |
| Maximum 02 | T = 6.36 | 4 | 16 | 64 | R Posterior-Medial Frontal |
| Maximum 03 | T = 5.49 | 8 | 12 | 62 | R Posterior-Medial Frontal |
| Maximum 04 | T = 5.41 | 2 | 12 | 70 | R Posterior-Medial Frontal |
| Maximum 05 | T = 5.27 | 10 | 16 | 58 | R Posterior-Medial Frontal |
| **Cluster 3 (29 vox): T- ( T> 4.79)** |  | | | | |
| Maximum 01 | T = 6.12 | 46 | -4 | -36 | R Inferior Temporal Gyrus |
| Maximum 02 | T = 5.98 | 42 | 0 | -34 | R Inferior Temporal Gyrus |
| Maximum 03 | T = 5.75 | 44 | -2 | -32 | R Inferior Temporal Gyrus |
| **Self-Location (F test)** | | | | | |
| **Cluster 1 (81 vox): F ( F> 29.25)** | **F/t** | **x** | **y** | **z** | **Area** |
| Maximum 01 | F = 104.76 | -58 | -48 | 28 | L Superior Temporal Gyrus |
| Maximum 02 | F = 83.64 | -54 | -48 | 30 | L SupraMarginal Gyrus |
| Maximum 03 | F = 80.97 | -50 | -46 | 32 | L SupraMarginal Gyrus |
| Maximum 04 | F = 52.26 | -48 | -46 | 28 | L Superior Temporal Gyrus |
| Maximum 05 | F = 44.16 | -50 | -46 | 24 | L Superior Temporal Gyrus |
| **Cluster 2 (44 vox): F ( F> 29.25)** |  | | | | |
| Maximum 01 | F = 161.47 | 20 | -34 | -10 | R ParaHippocampal Gyrus |
| Maximum 02 | F = 118.05 | 24 | -22 | -30 | N/A |
| Maximum 03 | F = 110.90 | 14 | -30 | -14 | N/A |
| Maximum 04 | F = 63.87 | 12 | -32 | -10 | N/A |
| Maximum 05 | F = 52.53 | 22 | -24 | -20 | R ParaHippocampal Gyrus |
| Maximum 06 | F = 49.54 | 22 | -28 | -16 | R ParaHippocampal Gyrus |
| Maximum 07 | F = 44.03 | 18 | -30 | -12 | R ParaHippocampal Gyrus |
| Maximum 08 | F = 43.51 | 16 | -20 | -24 | N/A |
| **Self-Location (positive T test: Synch < Asynch)** | | | | | |
| **Cluster 1 (80 vox): T+ ( T> 4.79)** | **F/t** | **x** | **y** | **z** | **Area** |
| Maximum 01 | T = 12.71 | 20 | -34 | -10 | R ParaHippocampal Gyrus |
| Maximum 02 | T = 10.86 | 24 | -22 | -30 | N/A |
| Maximum 03 | T = 10.53 | 14 | -30 | -14 | N/A |
| Maximum 04 | T = 8.75 | 24 | -20 | -26 | N/A |
| Maximum 05 | T = 7.99 | 12 | -32 | -10 | N/A |
| Maximum 06 | T = 7.25 | 22 | -24 | -20 | R ParaHippocampal Gyrus |
| Maximum 07 | T = 7.04 | 22 | -28 | -16 | R ParaHippocampal Gyrus |
| Maximum 08 | T = 6.64 | 18 | -30 | -12 | R ParaHippocampal Gyrus |
| Maximum 09 | T = 6.60 | 16 | -20 | -24 | N/A |
| Maximum 10 | T = 5.74 | 14 | -38 | -16 | R Cerebelum (III) |
| Maximum 11 | T = 5.66 | 18 | -34 | -18 | R Cerebelum (III) |
| **Cluster 2 (55 vox): T+ ( T> 4.79)** |  | | | | |
| Maximum 01 | T = 10.62 | -2 | -60 | -8 | Cerebellar Vermis (4/5) |
| Maximum 02 | T = 10.19 | -8 | -56 | -10 | L Cerebelum (IV-V) |
| Maximum 03 | T = 7.46 | 12 | -48 | -10 | R Cerebelum (IV-V) |
| Maximum 04 | T = 7.03 | 2 | -60 | -16 | Cerebellar Vermis (6) |
| Maximum 05 | T = 6.99 | -4 | -50 | -4 | L Cerebelum (IV-V) |
| Maximum 06 | T = 6.52 | 6 | -50 | -4 | Cerebellar Vermis (4/5) |
| Maximum 07 | T = 6.20 | 6 | -48 | -8 | Cerebellar Vermis (3) |
| Maximum 08 | T = 6.10 | 4 | -56 | -8 | Cerebellar Vermis (4/5) |
| Maximum 09 | T = 5.94 | -6 | -56 | -6 | L Cerebelum (IV-V) |
| Maximum 10 | T = 5.51 | 14 | -50 | -12 | R Cerebelum (IV-V) |
| **Cluster 3 (53 vox): T+ ( T> 4.79)** |  | | | | |
| Maximum 01 | T = 8.08 | 0 | -30 | -16 | Brainstem (midbrain/pons)* |
| Maximum 02 | T = 7.80 | 2 | -34 | -18 | Brainstem (midbrain/pons)* |
| Maximum 03 | T = 6.58 | 2 | -30 | -8 | Brainstem (midbrain/pons)* |
| Maximum 04 | T = 6.52 | 2 | -34 | -26 | Brainstem (midbrain/pons)* |
| Maximum 05 | T = 5.67 | 4 | -36 | -28 | Brainstem (midbrain/pons)* |
| Maximum 06 | T = 5.64 | 0 | -28 | -20 | Brainstem (midbrain/pons)* |
| Maximum 07 | T = 5.32 | 6 | -34 | -20 | Brainstem (midbrain/pons)* |
| Maximum 08 | T = 5.21 | 4 | -36 | -22 | Brainstem (midbrain/pons)* |
| **Cluster 4 (48 vox): T+ ( T> 4.79)** |  | | | | |
| Maximum 01 | T = 9.09 | 18 | -38 | -50 | N/A |
| Maximum 02 | T = 8.87 | 6 | -46 | -56 | N/A |
| Maximum 03 | T = 7.69 | 14 | -40 | -50 | N/A |
| Maximum 04 | T = 7.18 | 10 | -44 | -50 | N/A |
| Maximum 05 | T = 5.90 | 18 | -46 | -46 | R Cerebelum (IX) |
| Maximum 06 | T = 5.71 | 10 | -46 | -54 | N/A |
| Maximum 07 | T = 5.71 | 8 | -48 | -60 | N/A |
| Maximum 08 | T = 5.69 | 22 | -44 | -46 | R Cerebelum (VIII) |
| **Self-Location (negative T test: Asynch < Synch)** | | | | | |
| **Cluster 1 (108 vox): T- ( T> 4.79)** | **F/t** | **x** | **y** | **z** | **Area** |
| Maximum 01 | T = 10.24 | -58 | -48 | 28 | L Superior Temporal Gyrus |
| Maximum 02 | T = 9.15 | -54 | -48 | 30 | L SupraMarginal Gyrus |
| Maximum 03 | T = 9.00 | -50 | -46 | 32 | L SupraMarginal Gyrus |
| Maximum 04 | T = 7.23 | -48 | -46 | 28 | L Superior Temporal Gyrus |
| Maximum 05 | T = 6.65 | -50 | -46 | 24 | L Superior Temporal Gyrus |

**Table S4: Local maxima of significant clusters per contrast, localised according to the Anatomy toolbox (V2.2b; Eickhoff et al., 2005), in SPM12.** (L = left hemisphere, R = right hemisphere; x, y, z = co-ordinates of maximum activated voxel in standard MNI152 space, F / t = F / t stat at this voxel. Peaks are listed at p<0.05 FDR cluster corrected (cluster-forming threshold: p<0.001). *No label in Anatomy toolbox.
